# Altered Platelet-Megakaryocyte Endocytosis and Trafficking of Albumin and Fibrinogen in *RUNX1* Haplodeficiency

**DOI:** 10.1101/2023.10.23.23297335

**Authors:** Fabiola Del Carpio-Cano, Guangfen Mao, Lawrence E. Goldfinger, Jeremy Wurtzel, Liying Guan, Afaque Mohammad Alam, Kiwon Lee, Mortimer E. Poncz, A. Koneti Rao

## Abstract

Platelet α-granules have numerous proteins, some synthesized by megakaryocytes (MK) and others not synthesized but incorporated by endocytosis, an incompletely understood process in platelets/MK. Germline *RUNX1* haplodeficiency, referred to as familial platelet defect with predisposition to myeloid malignancies (FPDMM), is associated with thrombocytopenia, platelet dysfunction and granule deficiencies. In previous studies, we found that platelet albumin, fibrinogen and IgG levels were decreased in a FPDMM patient. We now show that platelet endocytosis of fluorescent-labeled albumin, fibrinogen and IgG is decreased in the patient and his daughter with FPDMM. In megakaryocytic human erythroleukemia (HEL) cells, siRNA *RUNX1* knockdown (KD) increased uptake of these proteins over 24 hours compared to control cells, with increases in caveolin-1 and flotillin-1 (two independent regulators of clathrin-independent endocytosis), LAMP2 (a lysosomal marker), RAB11 (a marker of recycling endosomes) and IFITM3. Caveolin-1 downregulation in RUNX1-deficient HEL cells abrogated the increased uptake of albumin, but not fibrinogen. Albumin, but not fibrinogen, partially colocalized with caveolin-1. *RUNX1* knockdown increased colocalization of albumin with flotillin and of fibrinogen with RAB11 suggesting altered trafficking of both. The increased albumin and fibrinogen uptake and levels of caveolin-1, flotillin-1, LAMP2 and IFITM3 were recapitulated by shRNA *RUNX1* knockdown in CD34^+^-derived MK. These studies provide the first evidence that in *RUNX1-*haplodeficiency platelet endocytosis of albumin and fibrinogen is impaired and that megakaryocytes have enhanced endocytosis with defective trafficking leading to loss of these proteins by distinct mechanisms. They provide new insights into mechanisms governing endocytosis and α-granule deficiencies in *RUNX1-*haplodeficiency.

**Key points:**

1. Platelet content and endocytosis of α-granule proteins, albumin, fibrinogen and IgG, are decreased in germline RUNX1 haplodeficiency.
2. In *RUNX1*-deficient HEL cells and primary MK endocytosis is enhanced with defective trafficking leading to decreased protein levels.

## Introduction

Endocytosis is a major mechanism by which cells, including platelets and megakaryocytes (MK), take up diverse molecules to traffic them to distinct intracellular membrane compartments ^1–5^. Platelet α-granules have numerous proteins; some, such as platelet factor (PF4) and Von Willebrand factor (VWF), are synthesized by MK, and others, such as fibrinogen, albumin, IgG and factor V, are not synthesized by human MK but incorporated into granules by endocytosis and endosomal trafficking ^1^. In MK, following endocytosis proteins sequentially traffic from early endosomes to multivesicular bodies to late endosomes, and then to α-granules or to recycling endosomes for extrusion from the cell or to lysosomes for degradation ^6^. Our understanding of mechanisms regulating endocytosis, trafficking and granule formation in MK and platelets is incomplete.

Transcription factor RUNX1 is a major regulator of definitive hematopoiesis, megakaryopoiesis, and platelet production ^7–9^. Human *RUNX1* haplodeficiency (RHD) due to heterozygous germline mutations is associated with familial thrombocytopenia, platelet dysfunction, α- and dense- granule (DG) deficiencies, and a predisposition to myeloid malignancies – a constellation termed familial platelet disorder with associated myeloid malignancy (FPDMM) ^10–13^. Platelet granules deficiencies are a hallmark of FPDMM ^11,14,15^ but the underlying mechanisms poorly understood.

In prior studies in a patient with FPDMM we described the presence of α- and dense granule deficiencies, and impaired activation induced aggregation, secretion, protein phosphorylation (pleckstrin and myosin light chain), and αIIbβ3 activation ^11,15–18^. On platelet expression profiling many genes were downregulated ^17^ and we have shown several to be direct RUNX1 targets ^14,19–21^, including *PF4* ^20^ and *MYL9* ^19^. Our recent studies revealed that *RAB31* ^22^ and *RAB1B* ^23^ – two small GTPases involved in endosomal trafficking (both *RUNX1* targets) are downregulated and associated with defective trafficking of VWF, mannose 6-phosphate receptor (M6PR) and epidermal growth factor (EGFR). An important finding in our patient was that platelets had decreased albumin, fibrinogen and IgG ^18^ - proteins not synthesized by MK but incorporated by endocytosis^1^. To understand the mechanisms, we pursued the hypothesis that endocytosis and trafficking are impaired in platelets/MK in *RUNX1-* deficiency. Our studies provide the first evidence that FPDMM platelets have defective endocytosis of albumin, fibrinogen, and IgG. In *RUNX1-*deficient megakaryocytic HEL cells and MK differentiated from human CD34^+^ cells, uptake of albumin and fibrinogen was increased with impaired trafficking by distinct mechanisms leading to decreased cellular levels. There was upregulation of caveolin-1 (Cav1) and flotillin-1 (Flot1, two proteins linked to clathrin-independent endocytosis ^4,24^, and of lysosomal marker LAMP2 ^25^ and recycling endosomal marker RAB11 ^25,26^, both linked to endosomal trafficking. These studies provide the first evidence that MK endocytosis and trafficking are perturbed in RHD.

## Methods

### Patient Information

The patient is a previously described ^(1–3)^ white male in his 40’s with FPDMM with a point mutation in *RUNX1* (*RUNX1* c.352-1 G>T), in intron 3 at the splice acceptor site for exon 4, leading to a frameshift with premature termination in the conserved Runt homology domain. We studied his daughter who has the same mutation. Healthy control subjects were recruited from staff and students at the Lewis Katz School of Medicine at Temple University. This research was approved by institutional Human Subjects Review Board and all participants gave a written informed consent.

### Reagents

Reagents were obtained from vendors as indicated: human fibronectin (EMD Millipore, Burlington MA), poly-L-lysine solution (SIGMA Life Science, St. Louis, MO), SPHERO Ultra Rainbow fluorescent particles (Spherotech, Inc. Lake Forest, IL), phorbol 12-myristate 13-acetate (PMA) (Enzo Life Sciences, Farmingdale, NY). Human fibrinogen (Haematologic Technologies, Essex Junction, VT). Pitstop 2 (Cayman Chemical, Ann Arbor). siRNAs against *RUNX1*, *CAV1*, *FLOT1, IFITM3,* and *CLTC* and control siRNAs, were purchased from Santa Cruz Biotechnology Inc, (Dallas, Texas). Antibodies used are shown in Supplemental Table 1.

### Preparation of platelets

Platelet-rich plasma (PRP) was prepared by centrifugation (200g, 20 min) from whole blood collected in 1/10 volume of 3.8% sodium citrate and incubated with carbacyclin (30 nM) for 20 minutes at room temperature (RT). Platelets were pelleted by centrifugation (650g, 15 min, RT), resuspended in HEPES Tyrode’s buffer after washing twice by centrifugation (650g, 10 minutes, RT). and used in experiments.

### Studies in HEL cells

Human erythroleukemia (HEL) cells obtained from the American Type Culture Collection (Rockville, MD) were cultured in RPMI 1640 medium (Cellgro, Manassas VA), supplemented with 10% fetal bovine serum (FBS) (GE Healthcare, Mississauga) and penicillin/streptomycin (100 U/ml/100 mg/ml, Invitrogen) at 37°C in a humidified 5% CO_2_ atmosphere. Cells were treated with 30 nM phorbol myristate acetate to induce megakaryocytic transformation ^27^. PMA-treated HEL cells were transfected with 100 nM of *RUNX1, CAV1, FLOT1, IFITM3, CLTC* or control siRNAs using Lipofectamine reagents (Invitrogen, Carlsbad) and consisted of pools of three 20–25 bp oligonucleotides (Santa Cruz Biotechnology, Dallas, TX). Cells were harvested at 24-48 hours after transfection and used for uptake studies.

### Studies in primary megakaryocytes

Primary MK were grown in-vitro from human CD34^+^ hematopoietic stem cell progenitors (HSPCs) as described and validated for studies on *RUNX1*-deficiency ^28,29^. CD34^+^ cells were infected with shRUNX1 or shNT-lentiviruses^28,29^ and cells expressing mCherry (mCherry+) were sorted on day 4 and cultured until day 11-12 to obtain RUNX-deficient (shRX) and control (shNT) MK used for studies.

### Protein uptake by flow cytometry

Protein uptake and retention (up to 24 hours) was studied in washed platelets, PMA-treated HEL cells and CD34+ cell-derived MKs suspended in buffer using flow cytometry. Platelet suspensions (1×10^6^/mL) were incubated with fluorescence conjugated Cy3-albumin (10 μg/mL), Cy3-fibrinogen (50 μg/mL) or Cy3-IgG (20 μg/ml) at 37°C for different times. HEL cells or MK (1.5×10^5^) were incubated with fluorescent conjugated albumin-Alexa Fluor 488 (30 μg/mL), fibrinogen-Alexa Fluor 647 (10 μg/mL) or IgG-Alexa Fluor 488 (30 μg/ml) at 37°C. Cells were fixed with 2% paraformaldehyde for 15 min at RT. Uptake-retention was evaluated using a BD^TM^ LSRII flow cytometry (San Jose, CA) and analysed with FlowJo software, v10.5.3. The LSRII flow cytometer was calibrated using standards beads, SPHERO Ultra Rainbow Fluorescent Particles (Spherotech, Inc. Lake Forest, IL) on each experimental day. Where indicated, cells were incubated (15 minutes, RT) prior to uptake studies with 30 μM Pitstop 2 (Cayman Chemical, Ann Arbor, MI), an inhibitor of clathrin-mediated mechanisms ^4^.

### Protein uptake by immunofluorescence microscopy

To assess protein uptake in immobilized platelets, washed platelets (1.5×10^6^/mL) were immobilized on coverslips precoated with 2 μg/mL human fibronectin (EMD Millipore, Rockville, MD) for 1 hr at 37°C; unbound platelets were removed by washing with HEPES Tyrode’s buffer (pH 7.4) and coverslips were incubated with fluorescent conjugated albumin-Alexa Fluor 488, fibrinogen-Alexa Fluor 647 or IgG-Alexa Fluor 488 (Jackson ImmunoResearch. West Grove, PA) at 37°C for indicated times. The coverslips were rinsed with HEPES Tyrode’s buffer pH 7.4 and fixed with 2% PFA for 15 minutes. Anti-PF4 antibody was used to mark platelet α-granules. Epifluorescence microscope (EVOS FL Auto imaging) was used to assess uptake.

HEL cells or MK were seeded on poly-L-lysine -coated coverslips and permeabilized with 0.1% Triton X-100 before immunostaining with antibodies as described ^15^. The antibodies used are shown in Supplemental Table 1. Images were obtained on an EVOS microscope or Leica TCS SP5 confocal microscope, using a 63X/1.40 n.a. oil immersion objective at room temperature and EVOS or Leica imaging software, respectively. Post-acquisition processing and analysis was performed with Adobe Photoshop and ImageJ ^30^ and was limited to image cropping and brightness/ contrast adjustments applied to all pixels per image simultaneously.

### Studies on ***αΙΙΒβ3*** expression and activation, and myosin light chain phosphorylation

HEL cells (1×10^6^/mL) treated with control or *RUNX1* siRNA were incubated with FITC-mouse anti-human CD41a clone–HIP8 or FITC mouse anti-human PAC1 antibody (Supplemental Table 1) to evaluate surface expression of αΙΙΒβ3 complex in the resting state and following activation with ADP (50 μM) or thrombin (10 U/mL) (Millipore Sigma Billerica, MA). Cells were fixed and evaluated by flow cytometry.

### Immunoblotting of cell lysates

Cell lysates collected in M-Per Protein Extraction Reagent (Pierce-Thermo Scientific) with protease inhibitors (Enzo Life Sciences, Farmingdale, NY) were subjected to 10% or 12% SDS-polyacrylamide gel electrophoresis, transferred to 0.2 μm nitrocellulose membranes and probed with antibodies. Proteins were detected with IRDye-labeled secondary antibodies using Odyssey Infrared Imaging system (Li-Cor Biosciences).

### Statistical analysis

Results were expressed as mean ± SEM. Differences were compared using Student’s t-test or 1- and 2-way ANOVA, using the GraphPad Prism, version 8 (GraphPad Software) and considered significant at *P < 0.05*.

## Results

### Platelet uptake of albumin, fibrinogen, and IgG is decreased in FPDMM

The uptake of Cy3-chrome-albumin, Cy3-chrome-fibrinogen and Cy3-chrome IgG over 15-30 minutes was markedly decreased in platelets from the father and daughter with *FPDMM* compared to a healthy subject (Figure 1A). These proteins are incorporated into α-granules ^1^. PF4 is another α-granule protein, but synthesized by MK and a direct *RUNX1*-target ^20^. PF4 immunofluorescence in platelets immobilized on fibronectin was reduced in the father, consistent with α-granule deficiency, but not the daughter (Figure 1B). We studied albumin-Alexa Fluor 488 and fibrinogen-Alexa Fluor 546 uptake in platelets immobilized on fibronectin using immunofluorescence microscopy (Figure 1C, D). The uptake of albumin was decreased at 1 and 15 min in the father and daughter compared to the healthy subject (Figure 1C). At 1, 5 and 15 min the father’s platelets had decreased fibrinogen; many platelets entirely lacked fibrinogen while others had substantial uptake (Figure 1D). In the daughter the uptake appeared normal. (Figure 1C). FPDMM patients are heterozygous for the RUNX1 mutation and heterogeneity with respect to platelet findings even among same family members has been observed ^31,32^. Overall, these findings involving 3 proteins studied by 2 approaches indicate that platelet endocytosis is defective in these patients.

**Figure 1.**
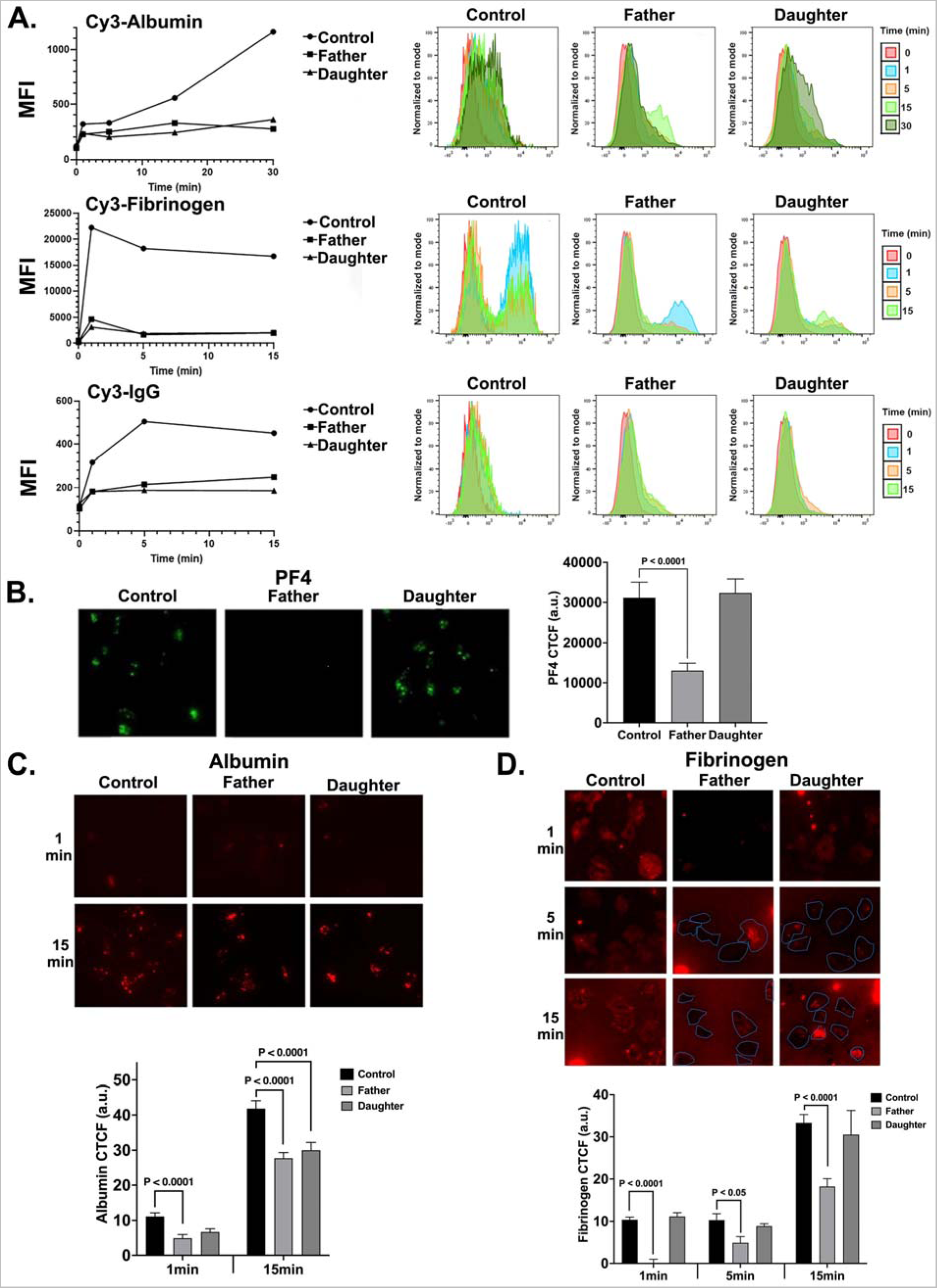
**A. Uptake of albumin, fibrinogen and IgG by platelets from father and daughter with FPDMM and a healthy control donor by flow cytometry.** Washed platelets (1×10^6^/mL) from the subjects were incubated with Cy3-albumin (10 μg/mL), Cy3- fibrinogen (50 μg/mL) or Cy3-IgG (20 μg/mL) for 15-30 minutes at 37°C. Platelets were fixed and analyzed by flow cytometry and expressed as mean fluorescent intensity (MFI). **B. Immunofluorescence studies of platelet** α**-granule PF4 in patients with FPDMM.** Platelets from the patients and a healthy control subject, immobilized on fibronectin-coated coverslips and fixed with 2% paraformaldehyde were incubated with FITC-labelled anti-PF4 monoclonal antibody and images were taken using Nikon E1000 microscope and quantified by image J (v1.47; National Institutes Health). Corrected total cell immunofluorescence (CTCF) is shown (mean ± SEM). The p values shown are for comparisons by Student’s t test. **C. Albumin uptake in platelets from patients with FPDMM immobilized on fibronectin and assessed by immunofluorescence microscopy.** Platelets from father, daughter and a healthy control subject immobilized on fibronectin-coated coverslips were incubated with albumin-Alexa Fluor 488 (30 μg/mL) (red) for the indicated time at 37°C and fixed. Images were taken with Nikon E1000 microscope to evaluate internalized albumin. The panel below shows the CTCF (mean±SEM) of albumin uptake in platelets from the patients and a control subject. The p values shown are for comparisons by Student’s t test. **D. Fibrinogen uptake in platelets from FPDMM patients immobilized on fibronectin and assessed by immunofluorescence microscopy.** Platelets from father and daughter with FPDMM and a healthy control subject (Control) were immobilized on fibronectin-coated coverslips and incubated with fibrinogen-Alexa 546(50 μg/mL) (red) for the indicated time at 37°C. Images were analyzed as described above. Platelets are shadowed in blue line. Many platelets from the father were devoid of fibrinogen with a population with substantial uptake. Daughter’s platelets appeared similar to control. The panel below shows changes in CTCF of fibrinogen uptake in platelets from FPDMM patients and control subject. The p values are for comparisons by the Student’s t test.

### Increased uptake and loss of albumin, and upregulation of caveolin-1 and flotillin-1 in HEL cells on *RUNX1* knockdown

We assessed endocytosis in *RUNX1-*deficient HEL cells. In preliminary studies the uptake of albumin-Alexa 488 over 24 hours in PMA-treated HEL cells was concentration- (10 to 100 μg/ml) and time-dependent over 24 hours. With siRNA *RUNX1* KD, albumin uptake over 24 hours was increased compared to control cells (Figure 2A). Cav1 and Flot1 are two major proteins involved in endocytosis ^2–5^. Cav1 regulates albumin uptake in several cell types ^33,34^. On immunoblotting, control HEL cells showed minimal Cav1, which increased markedly on *RUNX1* downregulation (Figure 2B). While Flot1 was detected in control cells, it increased further on *RUNX1* downregulation (Figure 2B). siRNA *CAV1* downregulation did not appear to decrease albumin uptake compared to control cells; however, it abrogated the increase in albumin uptake noted on *RUNX1* knockdown (Figure 2A). *FLOT1* knockdown did not alter albumin uptake compared to control cells, and in contrast to the findings with *CAV1* downregulation, did not abrogate the increase in albumin uptake noted on *RUNX1* downregulation (Figure 2C). Immunoblotting confirmed the expected decreases in the respective proteins on knockdown of *RUNX1, CAV1 and FLOT1,* and the increases in Cav1 and Flot1 on *RUNX1* downregulation (Figure 2B). *CAV1* downregulation did not affect Flot1 expression; and vice-versa (not shown). Thus, *RUNX1* downregulation increased albumin uptake along with increased Cav1 and Flot1; the increased uptake was Cav1-dependent.

**Figure 2.**
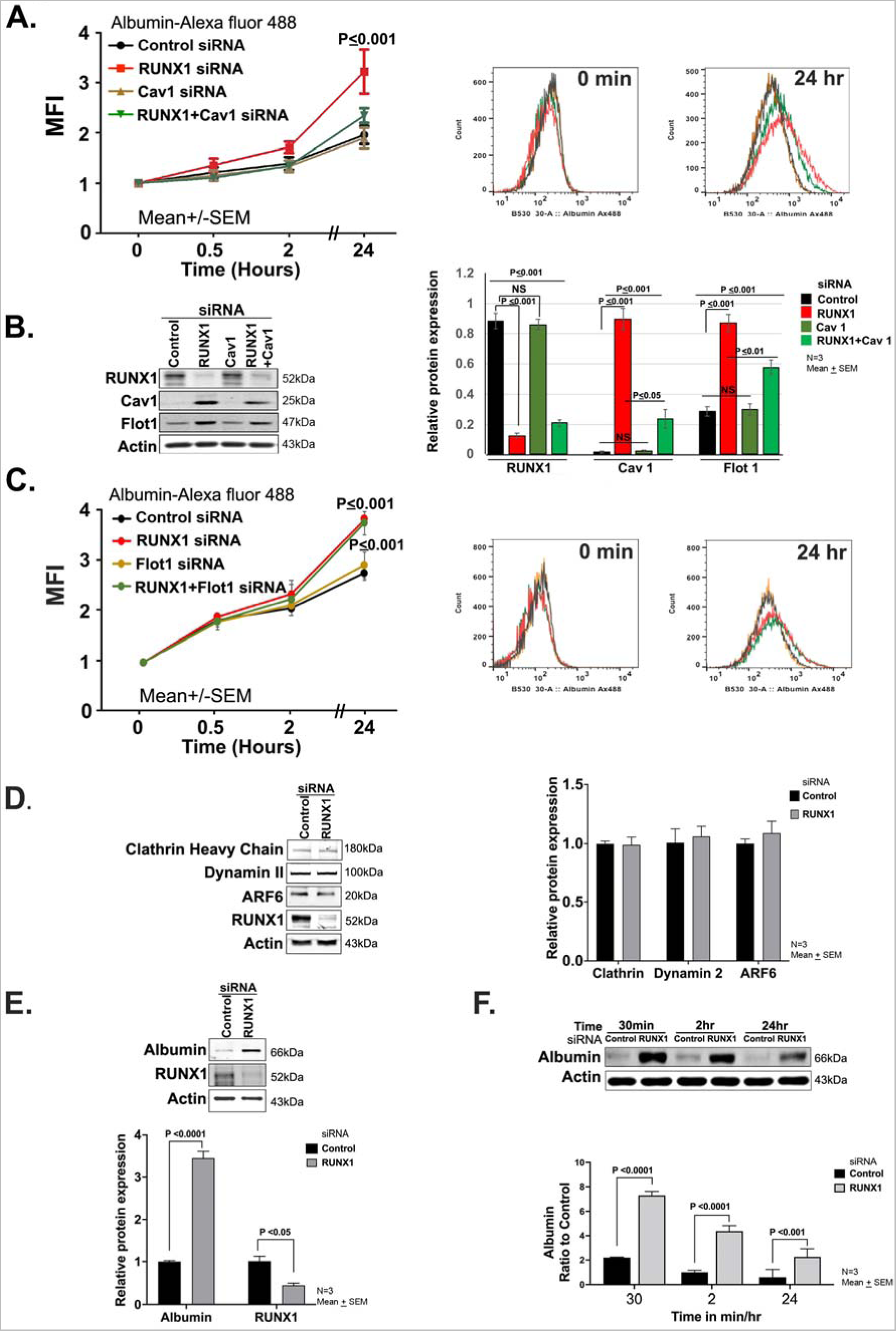
Effect of siRNA knockdown of *RUNX1*, *CAV1 and FLOT1* in HEL cells on uptake and retention of albumin. **A. Effect of siRNA knockdown of *RUNX1* and *CAV1* knockdown on uptake and retention of albumin by flow cytometry.** PMA-treated HEL cells were transfected with siRNA oligos (100 nM) targeting *RUNX1*, *CAV*1 or combination of both for 48 hours. They were incubated with 30 μg/mL albumin-Alexa 488 for the indicated period, fixed, washed, and albumin uptake was assessed by flow cytometry. MFI is shown (mean ± SEM of 3 experiments). Control siRNA (black line); *RUNX1* siRNA (red); *CAV1* siRNA (brown); *RUNX1* + CAV1 siRNA (green). P values shown are for comparisons with control siRNA by Student’s t test. **B.** Immunoblots showing RUNX1, Cav1, and Flot1 with actin as loading control in control cells and following KD of RUNX1, *CAV1,* or both. The quantification is shown on the right (mean±SEM; n=3). The p values are for comparisons by Student t test. **C.** Effect of siRNA knockdown of *RUNX1, FLOT1* or combination of both on uptake of albumin by flow cytometry. Following siRNA knockdown, HEL cells were incubated with 30 μg/mL of albumin-Alexa 488 for up to 24 hrs, fixed and analyzed by flow cytometry. Shown is albumin uptake expressed as MFI (mean±SEM, n=3). The p values shown are for comparisons with control siRNA by Student’s t test. Control siRNA (black), *RUNX1* siRNA (red), *FLOT1* siRNA (brown), *RUNX1+FLOT1* siRNA (green). **D.** Immunoblots showing relative levels of clathrin, dynamin-2 and ARF6 following *RUNX1* KD in HEL cells. The relative protein expression is shown on the right (n=3). **E.** HEL cell albumin levels by immunoblotting following siRNA *RUNX1* knockdown. HEL cells were incubated with *RUNX1* siRNA for 24 hours in culture media containing fetal bovine serum. At 24 hours the cells were washed in buffer and albumin levels assessed in cell lysates. Shown are a representative immunoblot with quantification of albumin and RUNX1 on the right (n=4 experiments). **F.** HEL-cell albumin levels over 24 hours in control cells and *RUNX1-*deficient cells. HEL cells were incubated with *RUNX1* or control siRNA for 24 hours in culture media containing FBS. At 24 hours, cells were washed and suspended in buffer without albumin. Albumin levels were assessed in cell lysates at intervals shown. Shown is a representative immunoblot and quantification (n=4 experiments).

We examined other major proteins implicated in endocytosis. Clathrin-mediated endocytosis (CME) regulates fibrinogen uptake ^1^ and is dependent on dynamin-2 ^3,5,35^, and GTPase Arf6 ^36^. No changes were noted in these on *RUNX1* downregulation (Figure 2D).

In above experiments HEL cells were cultured in an albumin-rich medium with FBS. We assessed albumin in lysates of washed HEL cells after 24-48 hours of siRNA *RUNX1* knockdown. Albumin was increased on *RUNX1* knockdown (Figure 2E). To assess the fate of endocytosed albumin (labelled and unlabelled from the culture medium), siRNA-treated HEL cells were resuspended in buffer without FBS and levels assessed at 30 min, and 2 and 24 hours in lysates; *RUNX1*-deficient cells had higher albumin compared to control cells (Figure 2F) at each time. Under both conditions, the 24-hour albumin was lower than at the beginning, indicating loss over time, particularly in RUNX1-deficient cells.

### *RUNX1* knockdown increases fibrinogen uptake and loss in HEL cells

In preliminary studies, HEL-cell fibrinogen uptake was dose- and time-dependent over 24 hours. Fibrinogen-Alexa Fluor 647 uptake over 24 hours was increased on *RUNX1* knockdown compared to control cells (Figure 3A). *CAV1* downregulation of (Figure 3A) did not affect fibrinogen uptake in control cells nor the increased uptake on *RUNX1* downregulation. *FLOT1* (Figure 3B) knockdown alone did not affect the uptake; but appeared to partly blunt the increased uptake with RUNX1 knockdown. Thus, as with albumin (Figure 2), *RUNX1* knockdown increased fibrinogen uptake but in contrast, fibrinogen uptake was unaffected by *CAV1* knockdown.

**Figure 3.**
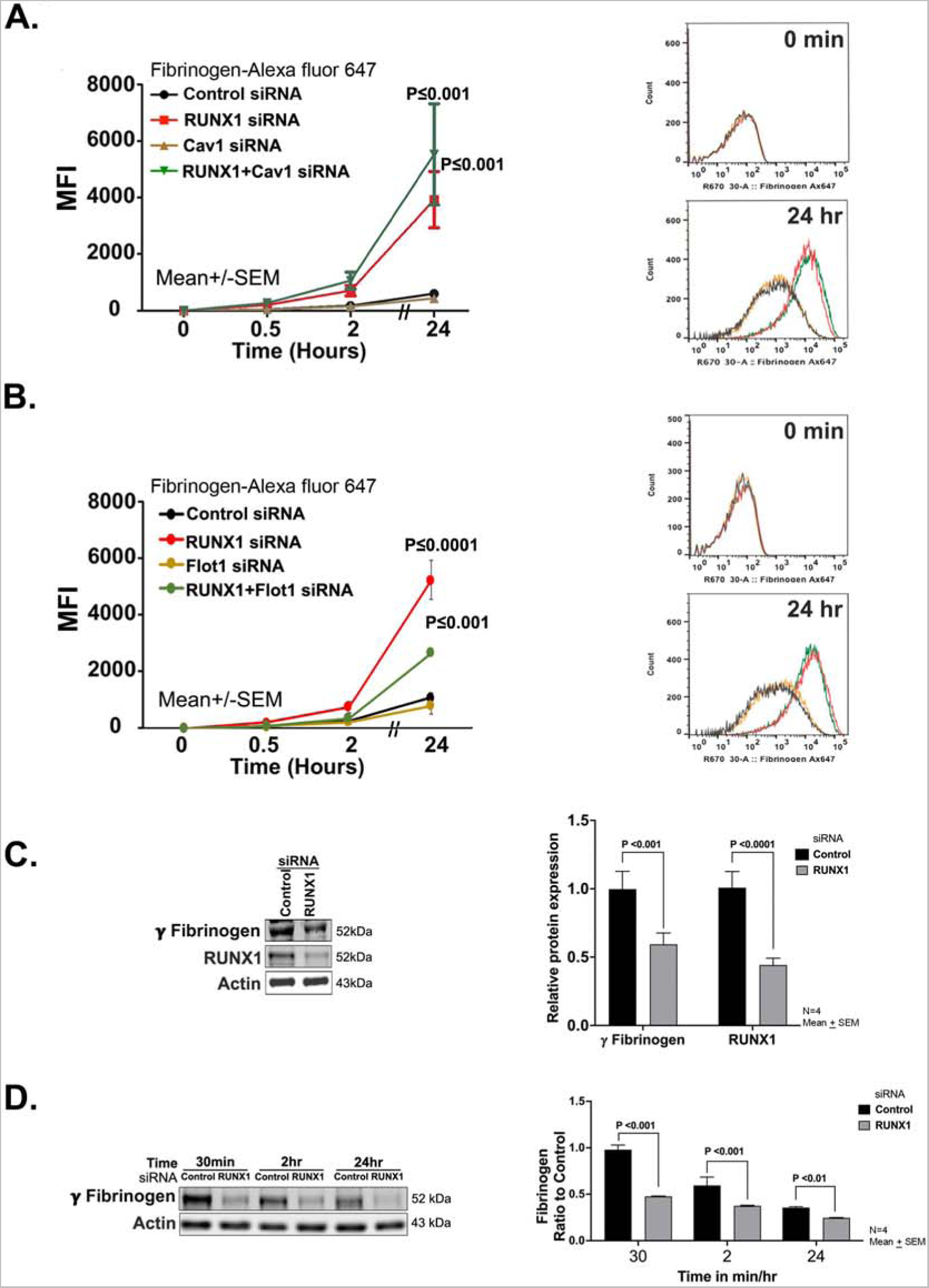
Effect of siRNA knockdown of *RUNX1*, *CAV1 and FLOT1* on uptake and retention of fibrinogen in HEL cells. **A.** Effect of siRNA KD of *RUNX1*, *CAV1* or combination of both on uptake and retention of fibrinogen. PMA-treated HEL cells were transfected with siRNAs (100 nM): control (black dots), *RUNX1* (red), *CAV1* (brown) or combination of *RUNX1* + *CAV1* (green) for 48 hours. Cells suspensions were incubated with 10 μg/mL fibrinogen-Alexa 647 for indicated times at 37°C, fixed and uptake was evaluated by flow cytometry. Shown mean ± SEM of 3 experiments. P values represent comparisons with control at 24 hours. **B.** Effect of siRNA knockdown of *RUNX1*, *FLOT1* or combination of both on uptake and retention of fibrinogen by flow cytometry. *RUNX1* and *FLOT1* knockdowns was performed as described above. HEL cells were incubated with fibrinogen-Alexa 647 (10 μg/mL) for indicated times. Data shown are mean of 2 experiments. Control cells (black), *RUNX1* KD (red), *FLOT1* KD alone (brown), combined *RUNX1* and *FLOT1* KD (green). **C.** HEL cell fibrinogen levels by immunoblotting following siRNA *RUNX1* knockdown. Cells treated with control and *RUNX1* siRNAs were washed and incubated in culture media containing FBS and 50 μg/ml fibrinogen for 24 hours. Fibrinogen levels were assessed in cell lysates. Shown is a representative immunoblot showing fibrinogen and RUNX1 with quantification on the right (n=4 experiments). **D.** Fibrinogen levels over 24 hours in HEL cells treated with control or *RUNX1* siRNAs and monitored in buffer without fibrinogen. HEL cells treated with *RUNX1* or control siRNAs were washed and resuspended in medium with added 10% FBS and 50 μg/ml fibrinogen for 24 hrs at 37°C. Cells were then washed, resuspended in medium without fibrinogen or FBS. Fibrinogen levels were assessed in lysates by immunoblotting at time points shown. Shown is a representative immunoblot and quantification from 4 separate experiments.

To understand the fate of the endocytosed fibrinogen, we measured fibrinogen by immunoblotting in washed HEL cells after 24-hour incubation in medium containing FBS and fibrinogen (50 μg/mL); *RUNX1*-deficient cells had lower levels compared to control cells (Figure 3C). In cells maintained in buffer over subsequent 24 hours fibrinogen was lower in *RUNX1*-deficient cells at 30 min, and 2 and 24 hours (Figure 3D) indicating continued decrease in cellular fibrinogen, despite enhanced endocytosis of the labelled protein at the plasma membrane.

### RUNX1 knockdown increases IgG uptake in HEL cells

Fluorescent conjugated IgG uptake was increased in *RUNX1*-deficient compared to control cells (Figure S1).

### Albumin colocalizes with caveolin-1 and this is lost with *RUNX1* knockdown and associated with increased colocalization with Flot1 and LAMP2

To understand the trafficking of albumin we performed immunofluorescence microscopy of HEL cells, stained for Cav1, Flot1, RAB11 - a marker for recycling endosomes ^25,26^, and LAMP2 - a marker for lysosomes ^25^. In control cells at 30 minutes, albumin colocalized with Cav1 but not with Flot1 (Figure 4A). On knockdown of *CAV1* alone or together with *RUNX1*, albumin was decreased indicating that *CAV1* regulates albumin uptake (Figure 4A). On *RUNX1* knockdown, there was loss of albumin colocalization with Cav1 and a strikingly increased colocalization with Flot1 (Figure 4A). There was no colocalization in the nuclear area (not shown). In control cells at 120 minutes albumin colocalization with Cav1 was not as strong as at 30 minutes and there was some albumin colocalization with Flot1 (Figure 4A). On *RUNX1* knockdown, there was little albumin colocalization with Cav1 and a strong colocalization with Flot1 (Figure 4A). With knockdown of *CAV1* alone or of *RUNX1*+*CAV1* there was no albumin in the cells. At 24 hours, albumin did not strongly colocalize with Cav1 or Flot1 in control or *RUNX1-*deficient cells (not shown). With respect to RAB11, immunoblots showed an increase on *RUNX1* knockdown (Figure 4B). However, at 30 and 120 minutes (Figure 4B) and at 24 hours (not shown), there was no albumin colocalization with RAB11 in control or *RUNX1-*deficient cells, suggesting that albumin does not traffic to recycling endosomes. In podocytes and other cells, albumin is degraded in lysosomes ^37,38^. LAMP2 levels were increased on *RUNX1* knockdown on immunoblotting (Figure 4C). We observed moderate colocalization of albumin with LAMP2 at 30 minutes in control cells and on *RUNX1* knockdown (Figure 4C) but they did not appear to be different. At 120 min the colocalization of albumin with LAMP2 appeared more prominent with *RUNX1* knockdown than in control cells (Figure 4C). Overall, RHD upregulates Cav1 and Flot1 (Figure 2), with altered albumin trafficking from Cav1- to Flot1- compartment, and increased colocalization with the lysosomal marker LAMP2.

**Figure 4.**
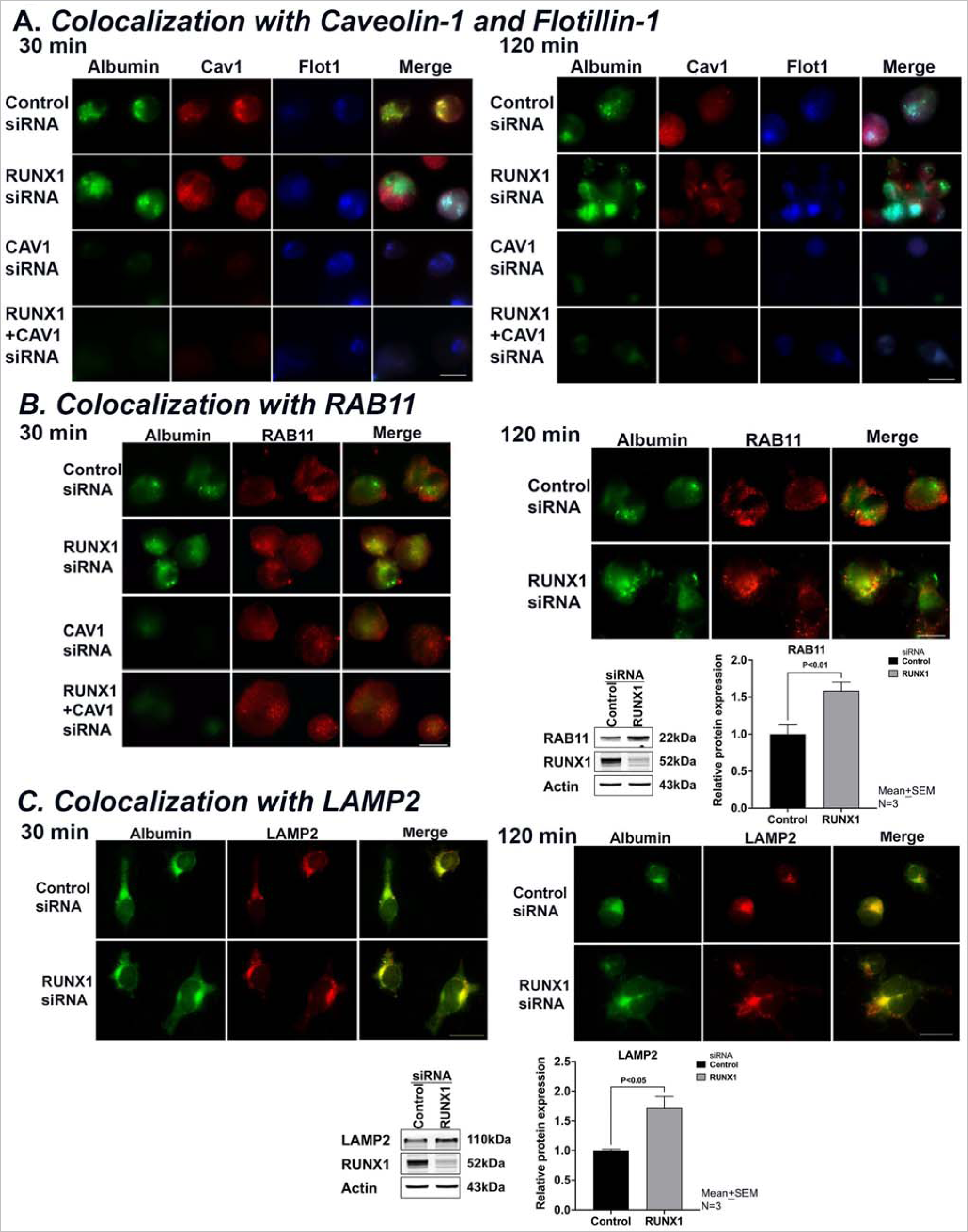
Effect of siRNA knockdown of RUNX1, CAV1 or both on albumin colocalization with Cav1, Flot1, RAB11 and LAMP2 in HEL cells by immunofluorescence microscopy. **A.** Representative images showing the effect of KD of *RUNX-1* or *CAV1* alone or in combination on albumin colocalization with Cav1 and Flot1. Control cells and those with siRNA knockdown of *RUNX1, CAV1* or the combination were incubated with 30 μg/ml albumin-Alexa 488 for 30 and 120 min, fixed and immobilized on poly-lysine-coated coverslips. Albumin is shown in green fluorescence. HEL cells were additionally stained with anti-Cav-1 (red) or anti-flotillin-1 (blue) antibodies to assess colocalization as seen in merged images, evaluated by Nikon E1000 epifluorescence microscope. At 30 min, in control cells albumin (green) was colocalized with Cav1 (red), as shown in yellow in merged images, but not with Flot1 (blue). With *RUNX1* KD, albumin colocalization with Cav1 was decreased with increased colocalization with Flot1. With C*AV1* KD alone no albumin was discernible. At 120 minutes, with *RUNX1* KD there was a decrease in albumin colocalization with Cav1 and an increase with Flot1. **B.** Representative images showing the effect of KD of *RUNX-1* or *CAV1* alone or in combination on albumin (green) colocalization with RAB11 (red), a marker for recycling endosomes. At 30 or 120 minutes, there was low colocalization of albumin (green) with RAB11 (red) in control cells or on *RUNX-1* KD. On *CAV1* KD alone or together with *RUNX1* KD there was negligible albumin in the cells. Representative immunoblots showing RAB11, RUNX1 and actin levels and densitometric quantification are shown. Data is shown as means ± SEM (n=4). **C.** Representative images showing the effect of knockdown of *RUNX-1* or *CAV1* alone or in combination on albumin (green) colocalization with lysosomal marker LAMP2 (red). At 30 minutes there was some colocalization of albumin with LAMP2 in control cells and on *RUNX-1* KD. At 120 min the colocalization of albumin with LAMP2 appeared more prominent with *RUNX1* KD than in control cells. Representative immunoblot showing protein expression of LAMP2, *RUNX1* and actin, and densitometric quantification. Shown as mean ± SEM (n=4).

### Increased colocalization of fibrinogen with RAB11 on *RUNX1* knockdown

Fibrinogen uptake was unaffected by *CAV1* knockdown (Figure 5A) suggesting that it is not Cav1-dependent. In control cells or on *RUNX1* knockdown, we did not observe significant fibrinogen colocalization with Cav1 or Flot1 at 30 or 120 min (Figure 5A). Findings with respect to RAB11 were different. At 30 minutes we observed low to moderate fibrinogen colocalization with RAB11 in control cells, which increased with *RUNX1* knockdown alone or together with *CAV1* (Figure 5B); colocalization with RAB11 was more prominent at 120 min (Figure 5B) and 24 hours (not shown). As noted earlier, RAB11 levels were increased with *RUNX1* downregulation (Figure 4B). These findings suggest that *RUNX1* knockdown causes trafficking of fibrinogen towards recycling endosomes. In both control cells and *RUNX1*-depleted cells we observed moderate fibrinogen colocalization with LAMP2 at 30 min but was not different (**Figure 5C**).

**Figure 5.**
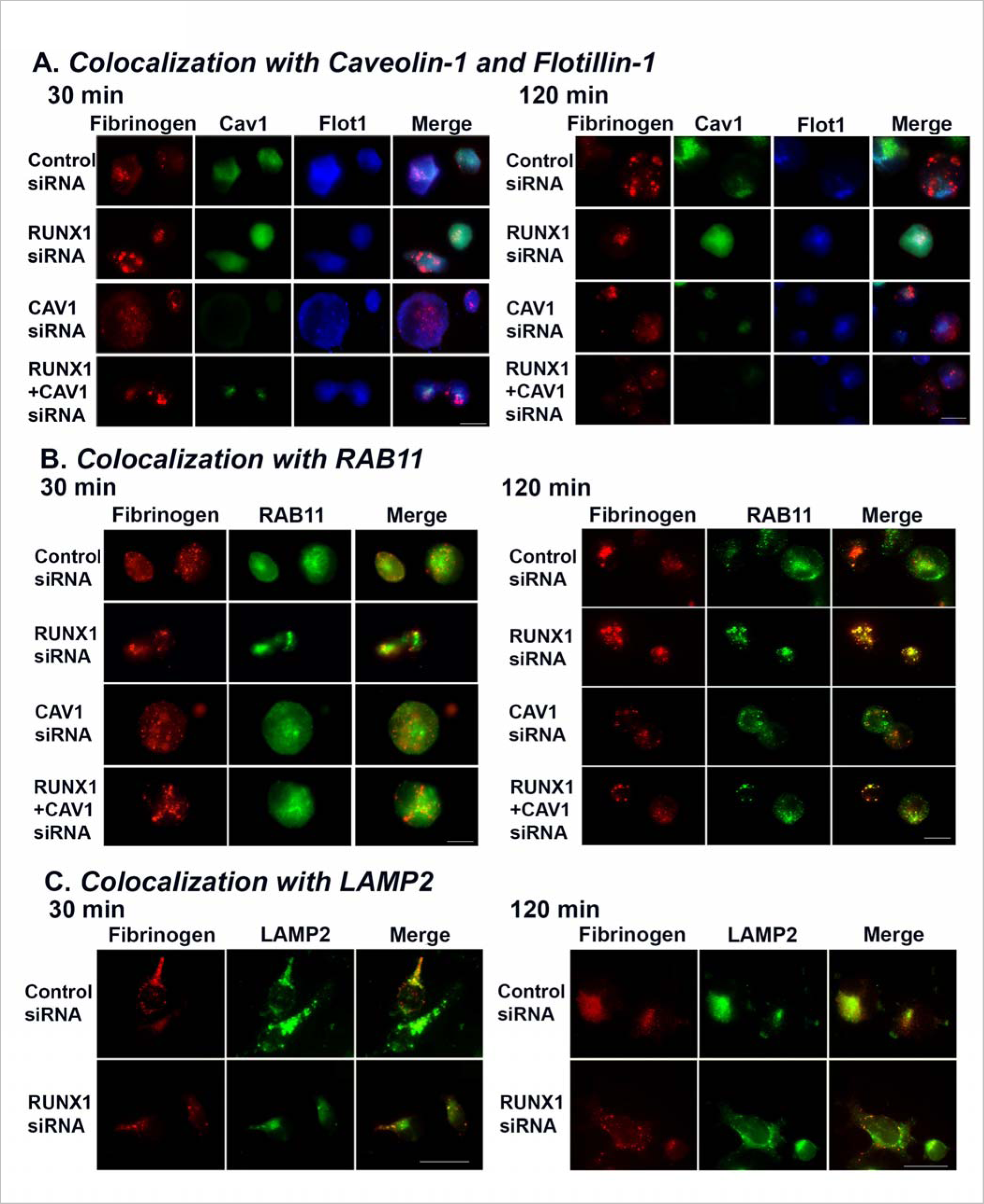
Effect of siRNA knockdown of RUNX1, CAV1 or both on fibrinogen colocalization with Cav-1, Flot1, RAB11 and LAMP2 in HEL cells by immunofluorescence microscopy. **A.** Representative images showing the effect of KD of *RUNX-1* or *CAV1* alone or in combination on fibrinogen colocalization with Cav1 and Flot1. The experimental design was same as in Figure 4. HEL cell suspensions (control cells and those with siRNA KD of *RUNX1, CAV1* or combination) were incubated with fibrinogen-Alexa 647, fixed and immobilized on poly-lysine-coated coverslips. HEL cells were additionally stained with anti-Cav-1 (green) or anti-flotillin-1 (blue) antibodies to assess colocalization as seen in merged images, evaluated by Nikon E1000 epifluorescence microscope. At 30 and 120 minutes, in control cells fibrinogen (red) showed no colocalization with Cav1 (green) or with Flot1 (blue). With C*AV1* KD alone fibrinogen uptake was unaffected compared to control cells. **B.** Representative images showing the effect of KD of *RUNX-1*, *CAV1* KD or the combination on fibrinogen (red) colocalization with RAB11 (green), a marker for recycling endosomes. At 30 minutes there was low to moderate fibrinogen colocalization (merged images, yellow) with RAB11 in control cells, and this was increased with *RUNX-1* KD and with KD of *RUNX1+CAV1* (left panel); the colocalization with RAB11 was more prominent at 120 min (right panel). **C.** Representative images showing the effect of KD of *RUNX-1* or *CAV1* KD alone or in combination on colocalization of fibrinogen with lysosomal marker LAMP2 (green). At 30 (left panel) or 120 (right panel) minutes, there was similar low colocalization of fibrinogen (red) with LAMP2 (green) in control cells or on *RUNX-1* KD.

### RUNX1*-*deficient HEL cells recapitulate the defective **α**IIb**β**3 activation and myosin phosphorylation in FPDMM platelets

Megakaryocyte fibrinogen uptake is αIIbβ3 receptor-mediated ^1,36,39,40^. Our patient platelets have normal surface αIIbβ3 expression but impaired agonist-induced activation (PAC binding) and MLC phosphorylation (pMLC) ^16^, latter related to decreased expression of RUNX1-regulated gene *MYL9* ^17^. Because fibrinogen uptake was increased on *RUNX1* knockdown (Figure 3) we assessed whether the HEL cells recapitulate these platelet abnormalities or show increased αIIbβ3. αIIbβ3 surface expression in HEL cells on *RUNX1* knockdown was comparable to that in control cells (Figure 6A); on immunoblotting αIIb was decreased with unchanged β3 (Figure 6B). Because the surface αIIbβ3 driving fibrinogen uptake is preserved (Figure 6A), the impact of the decreased cell lysate αIIb would be negligible. In HEL cells with *RUNX1* knockdown thrombin and ADP-induced PAC1-binding (Figure 6C), total MLC protein and, correspondingly, thrombin-induced pMLC (Figure 6D) were decreased. Thus, *RUNX1-*deficient HEL cells recapitulate the platelet defects ^16–18^, validating the HEL cell model. When pMLC was expressed as a fraction of the total myosin, it was comparable between RHD HEL cells and control cells (Figure 6D), suggesting that phosphorylation mechanisms (MLC kinase-driven) were preserved. These studies indicate that increased fibrinogen uptake is not due to upregulated αIIbβ3 expression. Because αIIbβ3 activation is impaired in these cells (Figure 6C), it suggests that integrin activation is not a major driver of fibrinogen uptake.

**Figure 6.**
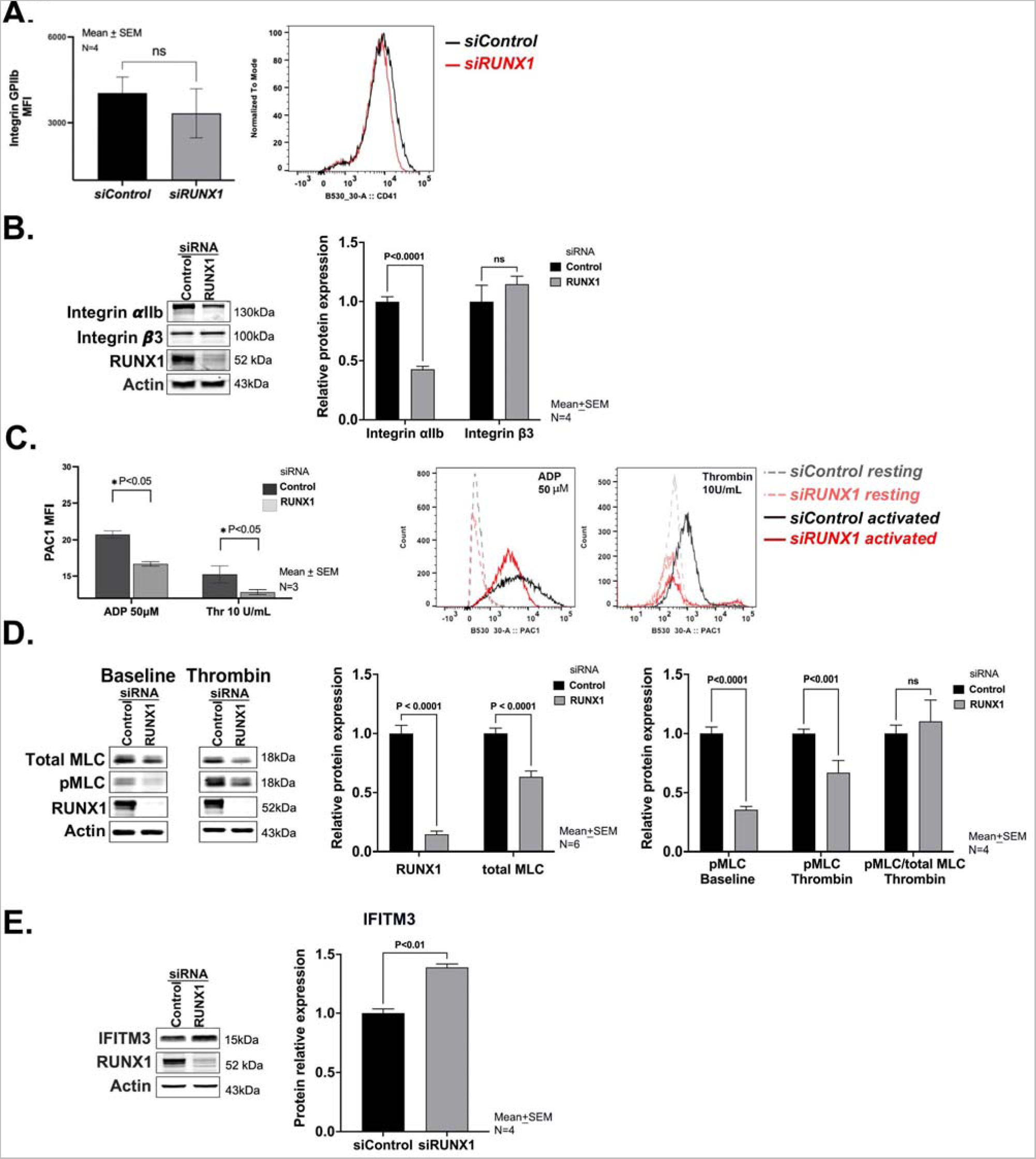
Effect of *RUNX1* KD on αIIbβ3 expression and activation, myosin light chain expression and phosphorylation, and IFITM3 in HEL cells. **A.** Effect of *RUNX1* KD on αIIbβ3 expression in HEL cells by flow cytometry using anti-CD41a antibody. **B.** Representative immunoblots of αIIb, β3, RUNX1 and β-actin protein expression and densitometric quantification (n=4). The p values show comparisons by the Student’s t test. **C.** Effect of *RUNX1* KD on αIIbβ3 activation evaluated by flow cytometry. FITC-labelled PAC1 antibody was used to assess activation of αIIbβ3 complex in control HEL cells (black lines) and following *RUNX1* KD (red lines), in the resting state (interrupted lines) and upon stimulation with ADP (50 μM) or thrombin (10 U/mL) (continuous lines). Data expressed in arbitrary units as means ± SEM (n=3). **D.** Effect of *RUNX1* KD on myosin light chain (MLC) expression and its phosphorylation upon thrombin activation in HEL cells. Left panel: Shown are representative immunoblots of total MLC, phospho-myosin light chain (pMLC), RUNX1 and actin protein expression in control cells and those with *RUNX1* KD, in the resting state (baseline) and following thrombin activation (5 U/ml). Right panel: shows the relative protein expression of RUNX1 and total MLC, and of pMLC in resting state (baseline) and following thrombin (5 U/ml) activation in control and *RUNX1*-deficient cells. The last 2 bars show the pMLC as a fraction of the total MLC present in control cells and *RUNX1* deficient cells. **E.** Representative immunoblots showing relative protein expression in HEL cells of IFITM3, *RUNX1* and actin, and densitometric quantification on *RUNX1* knockdown. Data is shown as means ± SEM (n=4).

Because increased IFITM3 expression has been shown to enhance fibrinogen endocytosis in nonviral sepsis ^40^, we assessed levels in HEL cells; they were increased on *RUNX1* knockdown (Figure 6E). However, *IFITM3* knockdown did not decrease fibrinogen uptake in control or *RUNX1-*deficient cells (Figure S2A). Fibrinogen uptake is clathrin-dependent ^1,40^ and it was inhibited by clathrin inhibitor Pitstop 2 (Figure S2B).

### RUNX1-deficient primary megakaryocytes recapitulate enhanced endocytosis

Albumin (Figure 7A) and fibrinogen (Figure 7B) uptake at 24 hours was increased in shRX compared to control shNT-MK along with increased Cav1, Flot1, LAMP2 and IFITM3 (Figure 7C); RAB11 was decreased (Figure 7C). On immunofluorescence microscopy, there was little albumin colocalization with Cav1 in shNT or shRX MK. In shRX*-*MK there was increased albumin colocalization with Flot1 (30 and 120 minutes) and LAMP2 at 30 minutes compared to shNT-MK (Figure 7D). There was minimal fibrinogen colocalization with RAB11 or LAMP2 in shNT-MK (Figure S3). In shRX-MK, there was no increased fibrinogen colocalization with RAB11, as noted in HEL cells (Figure 4B). But RAB11 was decreased in shRX MK (Figure 7C) in contrast to an increase in RUNX1-deficient HEL cells (Figure 4B). Fibrinogen colocalization with LAMP2 appeared minimally increased at 120 minutes in shRX*-*MK (Figure S3). Further studies, beyond the scope of present studies, are needed to further delineate alterations in trafficking. The differences from the findings in HEL cells may reflect differences in cell maturity. Overall, shRX-MK recapitulate the enhanced protein uptake, increases in Cav1, Flot1, LAMP2 and IFITM3, and increased albumin colocalization with Flot1 and LAMP2.

**Figure 7.**
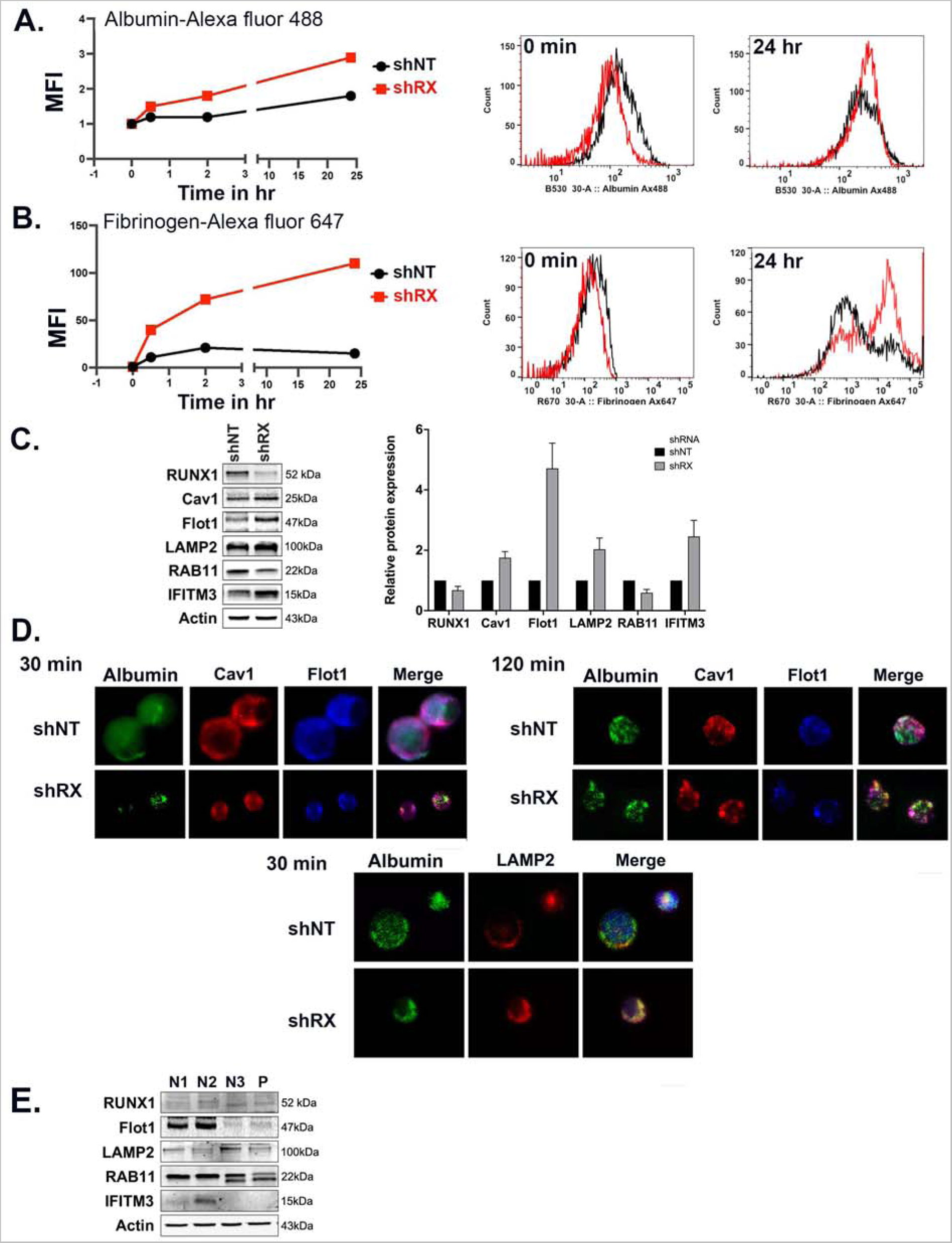
Effect of shRNA *RUNX1* knockdown in primary human megakaryocytes on albumin and fibrinogen uptake and platelet levels of flotillin-1, LAMP2, RAB11 and IFITM3 in FPDMM patient. **A.** Effect of shRNA *RUNX1* knockdown in primary MK on uptake of albumin by flow cytometry. MK were differentiated *in-vitro* from human CD34^+^ cells for 12 days. CD34^+^ cells were infected with sh*RX1-* or shNT-lentiviruses and expressing mCherry (mCherry+) and sorted on day 4 of differentiation. From day 5 cells were cultured until day 11 or day 12 to mature MK and used for protein uptake studies using flow cytometry as described for HEL cells. Left panel: line graph shows mean fluorescent intensity (MFI) of albumin-Alexa 488 uptake by MK overtime. Black lines indicate shNT and red shRX MK. Shown mean of 2 experiments. Right panel: Representative histograms of albumin-Alexa 488 uptake at the initial time point and 24 hours. **B.** Effect of shRNA *RUNX1* knockdown in MK on uptake of fibrinogen-Alexa 647 by flow cytometry. Black lines indicate shNT and red shRX MK. **C.** Effect of shRNA *RUNX1* knockdown in MK on levels of RUNX1, Cav1, Flot1, LAMP2, RAB11, IFITM3, and actin (as loading control) by immunoblotting. Shown on the right is the relative protein levels done in duplicate. **D.** Effect of shRNA RUNX1 knockdown in MK on albumin colocalization with Cav1, and Flot1 (top panels, 30 and 120 minutes) and with LAMP2 (bottom panel, 30 min) by immunofluorescence microscopy. Representative images are shown. shNT and shRX cells were incubated with 30 μg/ml albumin-Alexa 488 for 30 and 120 min, fixed and immobilized on poly-lysine-coated coverslips. Albumin is shown in green fluorescence. In the top panels, cells were additionally stained with anti-Cav1 (red) or anti-Flot1 (blue) antibodies to assess colocalization as seen in merged images. In the bottom panel the cells were additionally stained with anti-LAMP2 (blue). **E.** Platelet levels of Flot1, LAMP2, RAB11 and IFITM3 in FPDMM patient (P, father) and three healthy subjects (N1-3).

### Platelet proteins in FPDMM

Compared to 3 healthy donors, Flot1, LAMP2 and RAB11 were not increased in FPDMM platelets (Figure 7E). Cav1 was undetectable in all subjects, as reported ^35^. IFITM3 was undetectable in patient and one healthy subject. On platelet transcript profiling of our patient these genes were not increased ^17^. Thus, *RUNX1*-deficient platelets and HEL cells/MKs (early megakaryocytic cells) differ both in protein expression and phenotype (endocytosis).

## Discussion

Endocytosis is the mechanism by which proteins not synthesized by MK are incorporated into α-granules ^1^. Platelet albumin, fibrinogen and IgG were decreased in our patient ^18^ (Figure 1). We provide the first evidence that endocytosis is defective in FPDMM platelets (Figure 1), constituting one explanation. In addition, we provide evidence that *RUNX1-*deficient HEL cells and primary MK have increased endocytosis but also defective trafficking of albumin and fibrinogen, by distinct mechanisms, leading to decreased intracellular levels. Multiple pathways drive endocytosis, mediated by clathrin, Cav-1 and Flot1, and others ^2–4,41^. Cav1 and Flot1, but not clathrin, were increased in *RUNX1*-deficient HEL cells (Figure 2B) and primary MK (Figure 7C). *CAV1* knockdown near eliminated intracellular albumin (Figure 4) and the increase in albumin uptake with *RUNX1* knockdown (Figure 2), indicating that Cav1 regulates albumin uptake, as shown in renal podocytes ^33^. In contrast *CAV1* knockdown did not reduce intracellular fibrinogen (Figure 5), and *CAV1 (*Figure 3) knockdown did not abrogate the increased fibrinogen uptake on *RUNX1* knockdown, indicating that distinct mechanisms mediate albumin and fibrinogen endocytosis. MK from induced-pluripotent stem cells (IPSCs) from a FPDMM patient have been reported to show increased uptake of Factor V ^42^, also not synthesized by human MK. Thus, RHD increases MK endocytosis of multiple proteins. Interestingly, in *NBEAL2*-null mouse (a model of the gray platelet syndrome) MK fibrinogen uptake was normal but associated with defective retention ^43^, indicating differences from RHD.

MK fibrinogen uptake is αIIbβ3 receptor-mediated ^1,36,40^ and clathrin- and ARF6- dependent ^1,36^. In RUNX1-deficient HEL cells there was no increase in surface αIIbβ3 (Figure 6), clathrin (Figure 2D) or ARF6 (Figure 2D). Studies in sepsis advance IFITM3 as a regulator of platelet/MK fibrinogen endocytosis ^40^. IFITM3 was upregulated in *RUNX1-*deficient HEL cells and primary MK (Figures 6E and 7C) but *IFITM3* knockdown did not inhibit fibrinogen uptake (Figure S2) suggesting that in these cells it may not IFITM3-driven. IFITM3 effect see in sepsis ^40^ may require other interferon-induced genes as well ^44,45^.

Our studies suggest that *RUNX1* deficiency alters intracellular trafficking of albumin and fibrinogen and in a differential manner. *RUNX1* knockdown decreased albumin colocalization with Cav1 and increased with Flot1 (Figure 4). Cav1 and Flot1 are distinct clathrin-independent mechanisms of endocytosis ^2,46^ and flotillin and caveolin do not colocalize ^2^. The cargo of Flot1 positive endosomes has been shown to be delivered to lysosomes ^47,48^ with Flot1 present on cytosolic face of lysosomes ^49,50^. Lysosomal marker LAMP2 was increased on *RUNX1* downregulation and albumin colocalized with LAMP2 (Figures 4 and 7). Albumin has been reported to traffic to lysosomes where it is degraded ^33,38^. Thus, mis-trafficking of albumin with lysosomal degradation is a mechanism in RHD-MK with decreasing levels over time and in platelet progeny.

In MK, following endocytosis fibrinogen sequentially traffics from early endosomes to multivesicular bodies to late endosomes, and then to α-granules or recycling endosomes ^43^. Fibrinogen trafficking is perturbed in *RUNX1* deficiency. In *RUNX1*- deficient cells RAB11 was upregulated (Figure 4B) with increased fibrinogen colocalization with RAB11 (Figure 5B), which regulates protein recycling at plasma membrane ^26,51^, and cell fibrinogen was decreased (Figure 3C). We postulate that there is enhanced fibrinogen recycling in RHD-MK, as described in *NBEAL2^-/-^* MK with defective α-granules ^6^. Lysosomal degradation is not excluded in RHD because of the elevated levels and role of LAMP2 and Flot1 in degradation ^46,52,53^. Flotillins promote RAB11-dependent cargo recycling, as well ^53,54^.

Our studies in primary MK support remarkably well the findings in HEL cells with respect to the changes with RHD - increased uptake of albumin and fibrinogen, upregulation of several involved proteins and some aspects of mis-trafficking (Figure 7). Additional studies are needed in MK to delineate these mechanisms.

The overall model (see visual abstract, Supplement) that emerges is that RUNX1- deficient MK have enhanced endocytosis of albumin and fibrinogen but associated with defective intracellular trafficking and loss of protein via lysosomal degradation or recycling, leading to decreased levels in MK, despite the enhanced endocytosis, and subsequently in platelet progeny. Additionally, platelet have impaired endocytosis. The impaired trafficking and loss are likely related to defective granule biogenesis in RHD. These are driven by simultaneous dysregulation of numerous RUNX1-regulated genes ^17,28,55^. We have shown that RUNX1-regulated GTPases, *RAB1B* ^23^ and *RAB31*^22^, are decreased in RHD platelets and MK and associated with altered endosomal trafficking of VWF, M6PR, and EGFR and a striking defect with enlarged early-endosomes ^22^. The protein mis-trafficking applies also to other α−granule proteins - VWF and PF4 - synthesized by MK. Moreover, PF4 is a direct RUNX1 target, which also contributes to low levels in RHD ^20^.

IFITMs are important effectors in innate immunity and cancer biology ^44,45^ and RUNX1 is a negative regulator of neutrophil cytokine production ^56,57^. The IFITM3 upregulation in *RUNX1*-deficient HEL cells and MK (Figures 6 and 7) is novel and maybe relevant to autoimmune manifestations ^12,58^ and leukemic predisposition of FPDMM. RUNX1 may negatively regulate *IFITM3*. Platelet *IFITM3* mRNA levels are reported elevated in FPDMM ^55^. IFITM3 is implicated in the pathogenesis of malignancies ^44,45,59,60^ and AML patients with high expression have worse prognosis ^61^.

Our studies advance the complex mechanistic basis of the FPDMM abnormalities in α-granules and their cargos. These include dysregulated endocytosis and vesicular trafficking - poorly understood in platelet/MK but shared cellular processes that may be relevant to FPDMM manifestations beyond defective hemostasis.

## Data Availability

All data produced in the present study are available upon reasonable request to the authors

## Acknowledgements

We are grateful to the patients who have supported our studies over many years. This study was supported by research funding from NIH (NHLBI), R01 HL137376 and R01 HL109568 to AKR, and R01 HL137207 and HL159006 to LEG and R35 HL150698 to MP, American Heart Association Transformational Project Award 20TPA35490278 to LEG, and a RUNX1 Research Program/Alex’s Lemonade Foundation grant to MP. We thank David E. Ambrose and Amir Yarmahmoodi for discussions and assistance in the flow cytometry studies.

## Author Contributions

FDC, GFM and LG performed the research, analyzed the data, and contributed to the writing of the manuscript; JW performed the research and analyzed the data with respect to the immunofluorescence studies; AMA performed the research; KL and MP generously provided the MK differentiated from the human CD34^+^ cells with RUNX1 knockdown and their expertise; AKR and LEG conceived and designed and performed the research and interpreted data. AKR and LEG wrote the manuscript. All authors contributed to the manuscript, have read and approved the manuscript.

## References

1. Banerjee M, Whiteheart SW. The ins and outs of endocytic trafficking in platelet functions. Curr Opin Hematol. 2017;24(5):467–474.

2. Glebov OO, Bright NA, Nichols BJ. Flotillin-1 defines a clathrin-independent endocytic pathway in mammalian cells. Nat Cell Biol. 2006;8(1):46–54.

3. Parton RG. Caveolae: Structure, Function, and Relationship to Disease. Annu Rev Cell Dev Biol. 2018;34:111–136.

4. Rennick JJ, Johnston APR, Parton RG. Key principles and methods for studying the endocytosis of biological and nanoparticle therapeutics. Nat Nanotechnol. 2021;16(3):266–276.

5. Kaksonen M, Roux A. Mechanisms of clathrin-mediated endocytosis. Nat Rev Mol Cell Biol. 2018;19(5):313–326.

6. Kahr WH, Hinckley J, Li L, et al. Mutations in NBEAL2, encoding a BEACH protein, cause gray platelet syndrome. Nat Genet. 2011;43(8):738–740.

7. de Bruijn M, Dzierzak E. Runx transcription factors in the development and function of the definitive hematopoietic system. Blood. 2017;129(15):2061–2069.

8. de Bruijn MF, Speck NA. Core-binding factors in hematopoiesis and immune function. Oncogene. 2004;23(24):4238–4248.

9. Bonifer C, Levantini E, Kouskoff V, Lacaud G. Runx1 Structure and Function in Blood Cell Development. Adv Exp Med Biol. 2017;962:65–81.

10. Song WJ, Sullivan MG, Legare RD, et al. Haploinsufficiency of CBFA2 causes familial thrombocytopenia with propensity to develop acute myelogenous leukaemia. Nat Genet. 1999;23(2):166–175.

11. Songdej N, Rao AK. Hematopoietic transcription factor mutations: important players in inherited platelet defects. Blood. 2017;129(21):2873–2881.

12. Brown AL, Arts P, Carmichael CL, et al. RUNX1-mutated families show phenotype heterogeneity and a somatic mutation profile unique to germline predisposed AML. Blood Adv. 2020;4(6):1131–1144.

13. Sood R, Kamikubo Y, Liu P. Role of RUNX1 in hematological malignancies. Blood. 2017;129(15):2070–2082.

14. Mao G, Songdej N, Voora D, et al. Transcription Factor RUNX1 Regulates Platelet PCTP (Phosphatidylcholine Transfer Protein): Implications for Cardiovascular Events: Differential Effects of RUNX1 Variants. Circulation. 2017;136(10):927–939.

15. Mao GF, Goldfinger LE, Fan DC, et al. Dysregulation of PLDN (pallidin) is a mechanism for platelet dense granule deficiency in RUNX1 haplodeficiency. J Thromb Haemost. 2017;15(4):792–801.

16. Gabbeta J, Yang X, Sun L, McLane MA, Niewiarowski S, Rao AK. Abnormal inside-out signal transduction-dependent activation of glycoprotein IIb-IIIa in a patient with impaired pleckstrin phosphorylation. Blood. 1996;87(4):1368–1376.

17. Sun L, Gorospe JR, Hoffman EP, Rao AK. Decreased platelet expression of myosin regulatory light chain polypeptide (MYL9) and other genes with platelet dysfunction and CBFA2/RUNX1 mutation: insights from platelet expression profiling. J Thromb Haemost. 2007;5(1):146–154.

18. Sun L, Mao G, Rao AK. Association of CBFA2 mutation with decreased platelet PKC-θ and impaired receptor-mediated activation of GPIIb-IIIa and pleckstrin phosphorylation: proteins regulated by CBFA2 play a role in GPIIb-IIIa activation. Blood. 2004;103(3):948–954.

19. Jalagadugula G, Mao G, Kaur G, Goldfinger LE, Dhanasekaran DN, Rao AK. Regulation of platelet myosin light chain (*MYL9*) by RUNX1: implications for thrombocytopenia and platelet dysfunction in *RUNX1* haplodeficiency. Blood. 2010;116(26):6037–6045.

20. Aneja K, Jalagadugula G, Mao G, Singh A, Rao AK. Mechanism of platelet factor 4 (PF4) deficiency with RUNX1 haplodeficiency: RUNX1 is a transcriptional regulator of *PF4*. J Thromb Haemost. 2011;9(2):383–391.

21. Jalagadugula G, Mao G, Kaur G, Dhanasekaran DN, Rao AK. Platelet protein kinase C-theta deficiency with human RUNX1 mutation: PRKCQ is a transcriptional target of RUNX1. Arterioscler Thromb Vasc Biol. 2011;31(4):921–927.

22. Jalagadugula G, Mao G, Goldfinger LE, et al. Defective RAB31-mediated megakaryocytic early endosomal trafficking of VWF, EGFR, and M6PR in RUNX1 deficiency. Blood Adv. 2022;6(17):5100–5112.

23. Jalagadugula G, Goldfinger LE, Mao G, Lambert MP, Rao AK. Defective RAB1B-related megakaryocytic ER-to-Golgi transport in RUNX1 haplodeficiency: impact on von Willebrand factor. Blood Adv. 2018;2(7):797–806.

24. Parton RG, Collins BM. The structure of caveolin finally takes shape. Sci Adv. 2022;8(19):eabq6985.

25. Stenmark H. Rab GTPases as coordinators of vesicle traffic. Nat Rev Mol Cell Biol. 2009;10(8):513–525.

26. Takahashi S, Kubo K, Waguri S, et al. Rab11 regulates exocytosis of recycling vesicles at the plasma membrane. J Cell Sci. 2012;125(Pt 17):4049–4057.

27. Jalagadugula G, Dhanasekaran DN, Kim S, Kunapuli SP, Rao AK. Early growth response transcription factor EGR-1 regulates Galphaq gene in megakaryocytic cells. J Thromb Haemost. 2006;4(12):2678–2686.

28. Estevez B, Borst S, Jarocha D, et al. RUNX-1 haploinsufficiency causes a marked deficiency of megakaryocyte-biased hematopoietic progenitor cells. Blood. 2021;137(19):2662–2675.

29. Lee K, Ahn HS, Estevez B, Poncz M. RUNX1-deficient human megakaryocytes demonstrate thrombopoietic and platelet half-life and functional defects. Blood. 2023;141(3):260–270.

30. Schneider CA, Rasband WS, Eliceiri KW. NIH Image to ImageJ: 25 years of image analysis. Nat Methods. 2012;9(7):671–675.

31. Rao AK, Poncz M. Defective acid hydrolase secretion in RUNX1 haplodeficiency: Evidence for a global platelet secretory defect. Haemophilia. 2017;23(5):784–792.

32. Lui P, Cunningham L, Merguerian M, et al. RUNX1 NIH STUDY Blood. 2023;2023 (In Press).

33. Dobrinskikh E, Okamura K, Kopp JB, Doctor RB, Blaine J. Human podocytes perform polarized, caveolae-dependent albumin endocytosis. Am J Physiol Renal Physiol. 2014;306(9):F941–951.

34. Schubert W, Frank PG, Razani B, Park DS, Chow CW, Lisanti MP. Caveolae-deficient endothelial cells show defects in the uptake and transport of albumin in vivo. J Biol Chem. 2001;276(52):48619–48622.

35. Eaton N, Drew C, Wieser J, Munday AD, Falet H. Dynamin 2 is required for GPVI signaling and platelet hemostatic function in mice. Haematologica. 2020;105(5):1414–1423.

36. Huang Y, Joshi S, Xiang B, et al. Arf6 controls platelet spreading and clot retraction via integrin alphaIIbbeta3 trafficking. Blood. 2016.

37. Carson JM, Okamura K, Wakashin H, et al. Podocytes degrade endocytosed albumin primarily in lysosomes. PLoS One. 2014;9(6):e99771.

38. Gekle M. Renal tubule albumin transport. Annu Rev Physiol. 2005;67:573–594.

39. Handagama P, Scarborough RM, Shuman MA, Bainton DF. Endocytosis of fibrinogen into megakaryocyte and platelet alpha-granules is mediated by alpha IIb beta 3 (glycoprotein IIb-IIIa). Blood. 1993;82(1):135–138.

40. Campbell RA, Manne BK, Banerjee M, et al. IFITM3 regulates fibrinogen endocytosis and platelet reactivity in nonviral sepsis. J Clin Invest. 2022;132(23).

41. Doherty GJ, McMahon HT. Mechanisms of endocytosis. Annu Rev Biochem. 2009;78:857–902.

42. Borst S, Nations CC, Klein JG, et al. Study of inherited thrombocytopenia resulting from mutations in ETV6 or RUNX1 using a human pluripotent stem cell model. Stem Cell Reports. 2021;16(6):1458–1467.

43. Lo RW, Li L, Leung R, Pluthero FG, Kahr WHA. NBEAL2 (Neurobeachin-Like 2) Is Required for Retention of Cargo Proteins by alpha-Granules During Their Production by Megakaryocytes. Arterioscler Thromb Vasc Biol. 2018;38(10):2435–2447.

44. Friedlova N, Zavadil Kokas F, Hupp TR, Vojtesek B, Nekulova M. IFITM protein regulation and functions: Far beyond the fight against viruses. Front Immunol. 2022;13:1042368.

45. Gomez-Herranz M, Taylor J, Sloan RD. IFITM proteins: Understanding their diverse roles in viral infection, cancer, and immunity. J Biol Chem. 2023;299(1):102741.

46. Singh J, Elhabashy H, Muthukottiappan P, et al. Cross-linking of the endolysosomal system reveals potential flotillin structures and cargo. Nat Commun. 2022;13(1):6212.

47. Fan W, Guo J, Gao B, et al. Flotillin-mediated endocytosis and ALIX-syntenin-1-mediated exocytosis protect the cell membrane from damage caused by necroptosis. Sci Signal. 2019;12(583).

48. Stuermer CA, Lang DM, Kirsch F, Wiechers M, Deininger SO, Plattner H. Glycosylphosphatidyl inositol-anchored proteins and fyn kinase assemble in noncaveolar plasma membrane microdomains defined by reggie-1 and -2. Mol Biol Cell. 2001;12(10):3031–3045.

49. Kaushik S, Massey AC, Cuervo AM. Lysosome membrane lipid microdomains: novel regulators of chaperone-mediated autophagy. EMBO J. 2006;25(17):3921–3933.

50. Kokubo H, Helms JB, Ohno-Iwashita Y, Shimada Y, Horikoshi Y, Yamaguchi H. Ultrastructural localization of flotillin-1 to cholesterol-rich membrane microdomains, rafts, in rat brain tissue. Brain Res. 2003;965(1-2):83–90.

51. Savina A, Vidal M, Colombo MI. The exosome pathway in K562 cells is regulated by Rab11. J Cell Sci. 2002;115(Pt 12):2505–2515.

52. Wei D, Zhan W, Gao Y, et al. RAB31 marks and controls an ESCRT-independent exosome pathway. Cell Res. 2021;31(2):157–177.

53. Redpath GMI, Ecker M, Kapoor-Kaushik N, et al. Flotillins promote T cell receptor sorting through a fast Rab5-Rab11 endocytic recycling axis. Nat Commun. 2019;10(1):4392.

54. Bodrikov V, Pauschert A, Kochlamazashvili G, Stuermer CAO. Reggie-1 and reggie-2 (flotillins) participate in Rab11a-dependent cargo trafficking, spine synapse formation and LTP-related AMPA receptor (GluA1) surface exposure in mouse hippocampal neurons. Exp Neurol. 2017;289:31–45.

55. Palma-Barqueros V, Bastida JM, Lopez Andreo MJ, et al. Platelet transcriptome analysis in patients with germline RUNX1 mutations. J Thromb Haemost. 2023;21(5):1352–1365.

56. Bellissimo DC, Chen CH, Zhu Q, et al. Runx1 negatively regulates inflammatory cytokine production by neutrophils in response to Toll-like receptor signaling. Blood Adv. 2020;4(6):1145–1158.

57. Zezulin AU, Ye D, Howell E, et al. RUNX1 is required in granulocyte-monocyte progenitors to attenuate inflammatory cytokine production by neutrophils. bioRxiv. 2023.

58. Sorrell A, Espenschied C, Wang W, et al. Hereditary leukemia due to rare RUNX1c splice variant (L472X) presents with eczematous phenotype. Int J Clin Med. 2012;3(7).

59. Rajapaksa US, Jin C, Dong T. Malignancy and IFITM3: Friend or Foe? Front Oncol. 2020;10:593245.

60. Lee J. Does IFITM3 link inflammation to tumorigenesis? BMB Rep. 2022;55(12):602–608.

61. Liu Y, Lu R, Cui W, et al. High IFITM3 expression predicts adverse prognosis in acute myeloid leukemia. Cancer Gene Ther. 2020;27(1-2):38–44.

