## Supplementary material for "Altered Platelet-Megakaryocyte Endocytosis and Trafficking of Albumin and Fibrinogen in *RUNX1* Haplodeficiency": Supplemental Tables and Figures_102023.pdf

**Supplemental Table 1. List of antibodies used for immunoblots and immunofluorescence studies:**

| Primary Antibody | Source | Reactivity | Supplier | Reference |
|---|---|---|---|---|
| **Albumin** | Monoclonal Rabbit | Human, mouse, rat, cow | Abcam | ab192603 |
| **ARF6** | Polyclonal Rabbit | Human, mouse, rat | Proteintech | 20225-1-AP |
| **Caveolin-1** | Polyclonal Rabbit | Human, rat, mouse | Cell Signaling | 3238S |
| **Clathrin heavy chain** | Monoclonal Mouse | Human, mouse, rat, bovine | Novus Biologicals | NB300-613 |
| **Dynamin-2** | Polyclonal Rabbit | Human, mouse, rat | Cell Signaling | 2342S |
| **PAC1 (FITC-labeled)** | Mouse anti-human | Human, mouse | BD Biosciences | 340507 |
| **Flotillin-1** | Polyclonal Rabbit | Human, rat, mouse | Cell Signaling | 3253S |
| **IFITM3** | Polyclonal Rabbit | Human, mouse, rat | Proteintech | 11714-1-AP |
| **Integrin αIIb -FITC labeled clone-HIP8** | Mouse anti-human | Human, mouse | BD Biosciences | 340929 |

| | | | | |
|---|---|---|---|---|
| **Integrin αIIb** | Monoclonal Mouse | Human, mouse | Santa Cruz Biotechnology | sc-365938 |
| **Integrin β3** | Monoclonal Rabbit | Human, mouse | Cell Signaling | 13166S |
| **IRDye- IgG labeled 680/800 RD Sec. antibody** | Donkey | Human, mouse | Li-Cor Biosciences | 926-68072 |
| **LAMP2 CD107b** | Polyclonal Mouse | Human | Biolegend | 354302 |
| **Myosin light chain** | Monoclonal Mouse | Human, mouse | Santa Cruz Biotechnology | sc-48414 |
| **PF4 (anti-CXCL4)** | Monoclonal Human | Human | R&D Systems | IC-7952F |
| **Phosphorylated - myosin light chain** | Polyclonal Rabbit | Human, mouse | Cell Signaling | 3674S |
| **Rab11A/B** | Monoclonal Rabbit | Human, rat, mouse | Cell Signaling | 5589 |
| **RUNX1** | Monoclonal Mouse | Human, rat, mouse | Santa Cruz Biotechnology | sc-365644 |
| **β-actin** | Polyclonal Mouse | Human, mouse, rat | Santa Cruz Biotechnology | sc-47778 |

**Supplemental Table 2. List of fluorescence labeled conjugated proteins or antibodies used for uptake studies:**

| PROTEIN | CONJUGATES | SUPPLIER | REFERENCE |
|---|---|---|---|
| **Human Albumin** | Cy3-ChromePure | Jackson ImmunoResearch Laboratories, Inc | 009-160-051 |
| **Human Albumin** | Alexa Fluor 488 | Jackson ImmunoResearch Laboratories, Inc | 009-540-051 |
| **Fibrinogen from Human plasma** | Alexa fluor 546 | Invitrogen/Thermo Fisher sci | F13192 |
| **Fibrinogen from Human plasma** | Alexa Fluor 647 | Invitrogen/Thermo Fisher sci | F35200 |
| **Human IgG (whole molecule)** | Cy3-ChromePure | Jackson ImmunoResearch Laboratories, Inc | 009-160-003 |
| **Fibrinogen from Human plasma** | Cy3- Fibrinogen | ABBY monoclonal | Bsm-1240M-Cy3 |
| **Human IgG (whole molecule)** | Alexa Fluor 488 | Jackson ImmunoResearch Laboratories, Inc | 009-000-003 |

#### SUPLEMENTAL FIGURES

### Figure S1

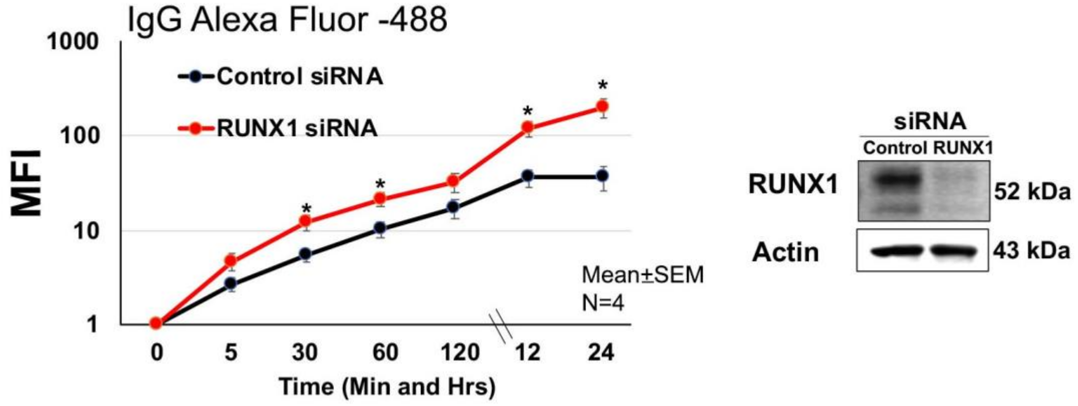


**Figure S1**. **Effect of *RUNX1* knockdown on IgG uptake in HEL cells.** HEL cells transfected with control or *RUNX1* siRNA were incubated with 10 μg/mL IgG-Alexa 488 at 37$^{o}$C for different time points, washed, fixed with 2% paraformaldehyde and analyzed by flow cytometry to assess IgG uptake. Shown is mean of MFI of two experiments. IgG uptake in *RUNX-1* deficient cells (red) was increased over time as compared to control cells (black). Also shown is a representative immunoblot showing *RUNX-1* expression with actin as loading control.

### Figure S2

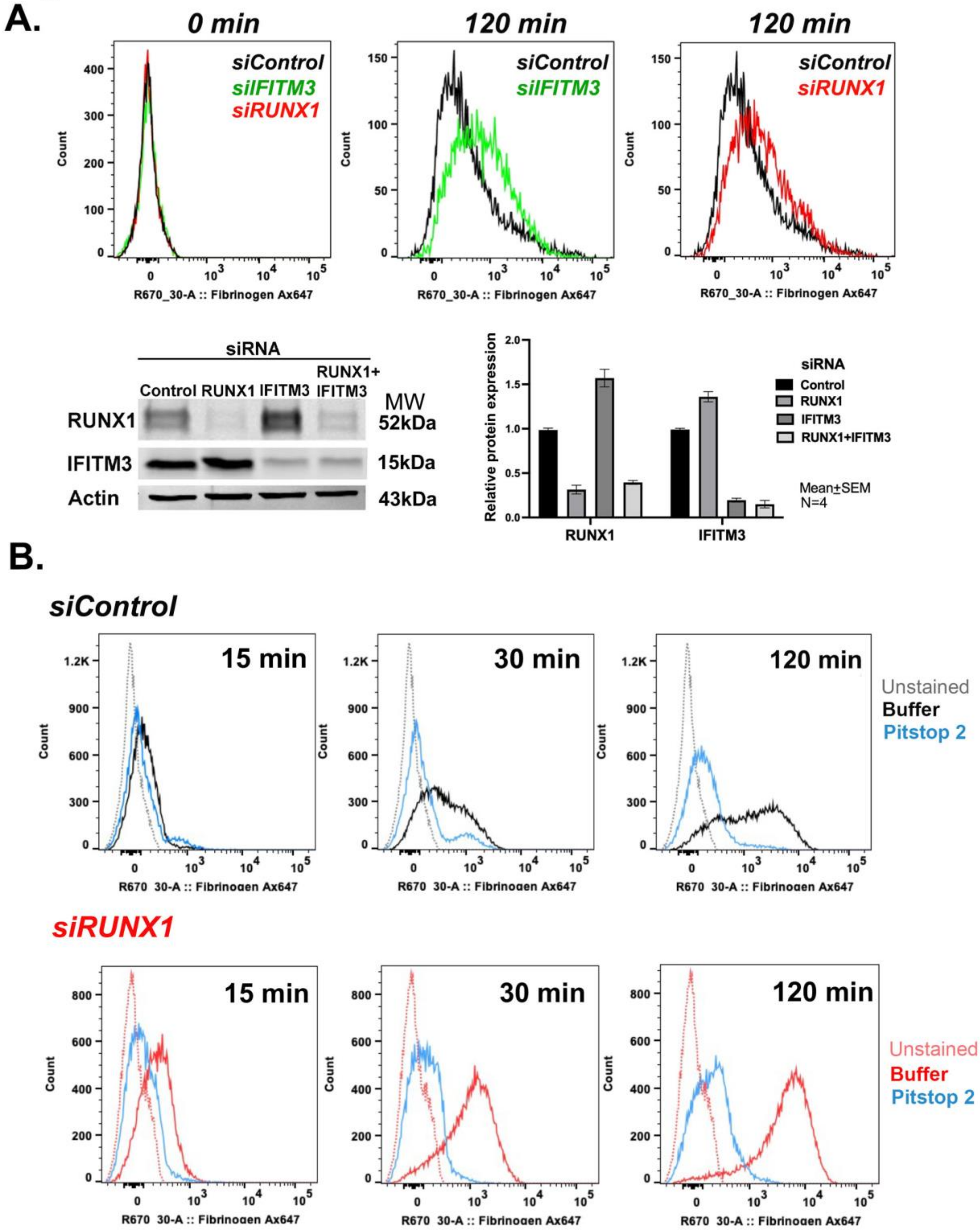


**Figure S2. Effect of *IFITM3* knockdown on fibrinogen uptake in HEL cells.**

**A.** Effect of knockdown of *IFITM3* or *RUNX1* on uptake of fibrinogen at 120 minutes by flow cytometry. HEL cells were transfected with siRNAs (100 nM) against *IFITM3* or *RUNX1*. Cells were incubated with 10 μg/mL Alexa 647-fibrinogen in buffer for 120 minutes at 37ºC, fixed and washed with buffer. Fibrinogen uptake was evaluated by flow cytometry. Left panel; 0 minutes. Middle panel: uptake at 120 minutes in *IFITM3 KD* cells (green) compared to control cells (black). The uptake of fibrinogen over 120 min was not inhibited on *IFITM3* knockdown. Right panel: uptake at 120 minutes in *RUNX1*-deficient cells (red) compared to control cells (black). Shown representative of 3 experiments. The immunoblots show the relative protein levels.

**B.** Effect of Pitstop 2 (inhibitor of clathrin-mediated uptake) on uptake and retention of fibrinogen over 120 min in control cells (top panels) and *RUNX1* siRNA treated HEL cells (lower panels) by flow cytometry. Cells were incubated with Pitstop 2 (30 μM) for 15 minutes at room temperature. In the top panels: black lines, buffer; blue lines Pitstop 2. In the lower panels: red lines, buffer; blue lines, Pitstop 2. Shown is a representative of 2 experiments.

#### Figure S3

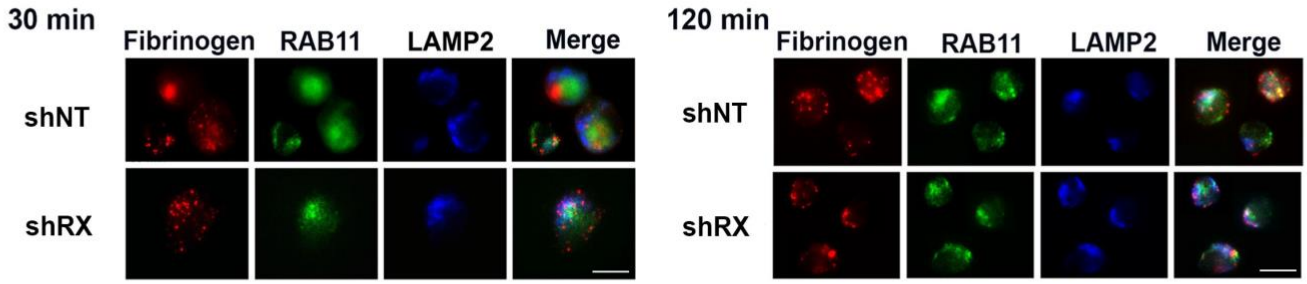


**Figure S3. Effect of shRNA RUNX1 knockdown in primary MK on fibrinogen colocalization with RAB11 and LAMP2 (30 and 120 minutes) by immunofluorescence microscopy.** shNT and shRx cells were incubated with Alexa-647 fibrinogen for 30 and 120 min, fixed and immobilized on poly-lysine-coated coverslips. Representative images are shown. Fibrinogen is shown in red fluorescence. Cells were stained with anti-RAB11 (green) or anti-LAMP2 (blue) antibodies to assess colocalization as seen in merged images and evaluated by Nikon E1000 epifluorescence microscope.

#### Figure S4

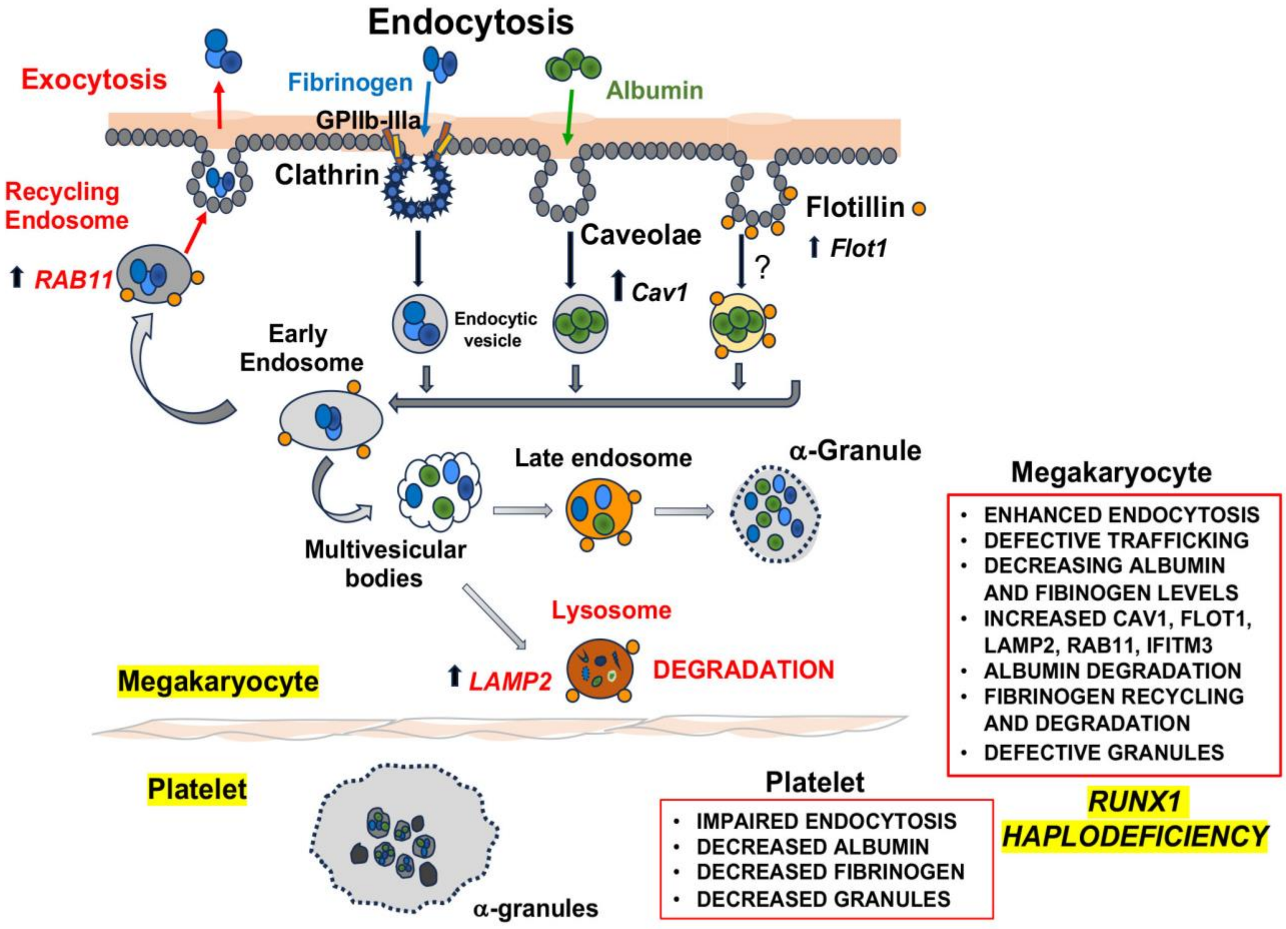


**Figure S4 Schema showing the endocytic pathways and alterations in megakaryocytes and platelets in *RUNX1* haplodeficiency.**
